# Use of Instagram and its effect on the Mental Well-being of University Students: A Pakistani Perspective

**DOI:** 10.1101/2025.11.11.25340027

**Authors:** Ayesha Qamar, Inayat ali

**Author notes:** Corresponding Author: Dr Inayat Ali Department of Anthropology, Fatima Jinnah Women University.

## Abstract

The use of social media has increased considerably in recent years. The impact of Instagram on university students in low-resource countries such as Pakistan requires further research. According to social media platforms, Pakistan had 54.38 million people aged 18 and older active on social media at the start of 2024. This figure accounts for 38.9 per cent of the total population aged 18 and above and prompted us to examine how Instagram use contributes to social comparison, leading to mental health issues, especially among university students. The study uses quantitative methods and online surveys to explore the relationship between Instagram use and mental well-being. A sample of 515 was recruited from two well-known universities in Islamabad through convenience sampling. The sample includes 515 male and female students aged between 18 and 25, applying a conditional mediation model (CoMe Model) evaluated by SmartPLS. The findings indicate that increased Instagram use strongly predicts decreased self-esteem (β = -0.661, p < .001), which is linked to higher levels of depression (β = -0.439, p < .001). The indirect effect of Instagram use on depression via self-esteem was significant (β = 0.290, p < .001), while the direct effect became non-significant when self-esteem was included, suggesting full mediation. Importantly, the strength of the mediated pathway varied with levels of upward comparison. The indirect effect was lower among those with high levels of upward comparison (β = 0.116) and stronger among those with low levels (β = 0.201), with the moderated mediation index also reaching significance (β = -0.035, p = .016). These results show that the psychological impact of Instagram use on mental health is variable and depends on users’ tendency to compare themselves with others.

## Introduction

Instagram has experienced substantial development as it ranks third among the most renowned social networking sites (1). Yet multiple studies have found a relationship between Instagram usage and mental well-being issues (2-5). Anxiety and depression have been associated with excessive social media usage (6). Instagram significantly impacted mental well-being, with 56.3% of participants reporting experiences of online hate on the platform, which led them to depression (7). In Pakistan, increased Instagram use among young girls substantially correlates with higher body dissatisfaction, mental any physical wellbeing (8). Likewise, Bint-e-Khalil and Ali (2025) have found that social medial impacts paramenters of beauty in women in Azad Jamu and Kashmir and affects their mental wellbeing (9). Individuals with more depressive symptoms compare themselves more upward on Instagram. Upward comparisons on Instagram increase depressive symptoms. The effects of depressive symptoms and social comparisons lead to a vicious circle (10, 11). Individuals with higher depressed symptoms are more driven to examine themselves, and engaging in upward comparisons on Instagram is connected with depression symptoms. Servidio, Soraci (12) indicated a significant link between fear of missing out (FoMO), social comparison, and problematic social media usage (PSMU). It was discovered that self-esteem and social comparison sequentially modulated the link between FoMO and PSMU.

This study focuses on examining the usage patterns of Instagram among university students in Pakistan and exploring how this social media platform impacts their mental well-being. Given the high youth population in Pakistan and the increasing popularity of Instagram, it is crucial to understand both the benefits and potential risks associated with its use. The study aims to identify specific factors related to Instagram that influence mental health, such as levels of social interaction, exposure to visual content, and online social support, while also considering cultural and socio-economic contexts unique to Pakistan. The present study pursues three primary objectives: (1) to investigate the association between Instagram usage and depression among university students in Pakistan; (2) to examine the mediating role of self-esteem in the relationship between Instagram usage and depression; and (3) to assess the moderating role of upward social comparison in the association between Instagram usage and depression among university students. To address these objectives, the study poses the following research questions: RQ1: What is the nature of the relationship between Instagram usage and depression among university students in Pakistan? RQ2: Does self-esteem mediate the relationship between Instagram usage and depression? RQ3: Does upward social comparison moderate the indirect effect of Instagram usage on depression via self-esteem? The central hypothesis (H1) posits that Instagram usage is significantly associated with depression among university students, with self-esteem acting as a mediator and upward social comparison serving as a moderator in this relationship.

### Proposed Conceptual Model of Research

## Methods and Materials

The research used quantitative analysis using SmartPLS 4 to assess the effect of social media usage and depression mediated by self-esteem and moderated by upward comparison (17). Kroenke, Spitzer (18) present a model that utilizes the Patient Health Questionnaire-9 (PHQ-9) to enhance depressive symptom identification on social media. Hamilton (19) adopted the questionnaire for measuring depression in young university students through the Patient Health Questionnaire-9PHQ-9. Upward comparison is measured by examining how SNS use (such as viewing Facebook profiles) affects upward social comparison, self-esteem, and subjective well-being (20, 21). A convenience sampling technique was used to collect the data from two universities in Islamabad (the capital of Pakistan). Bahria University comprises 10,000 students, of whom 8000 are undergraduate and 2000 are postgraduate students. Islamic International University IIUI has an enrollment of over 17,000 students, including around 7,000 students in undergraduate programs. The researcher selected 515 students, 260 males and 255 females, from undergraduate programs at both universities during 20 May 2025 to 20 July 2025.

The independent variable, usage of Instagram, is measured through 5 items, with responses ranging from “never” to “very often” = 5. In comparison, depression (PHQ-9) is measured through a 9-item scale, from not at all to nearly every day = 3. In contrast, the upward comparison is measured through a 5-item scale from strongly disagree=1 to strongly agree=5, and self-esteem is measured through a 10-item scale from strongly disagree=1 to strongly agree=5.

## Ethical Statement

The study adhered to the principles outlined in the Helsinki Declaration and was approved by the relevant institutional review board as per letter number: FJWU/EC/2025/105 on May 19, 2025. Furthermore, we obtained informed consent from all participants to participate and to have their data published. They had full opportunity and willingness to participate in this study or withdraw their participation at any stage. Except their views, no other specifimen were collected from them.

## Results

The study was conducted on the university students of two universities in Islamabad, Pakistan. The descriptive statistics are shown in Table 1

**Table 1:**
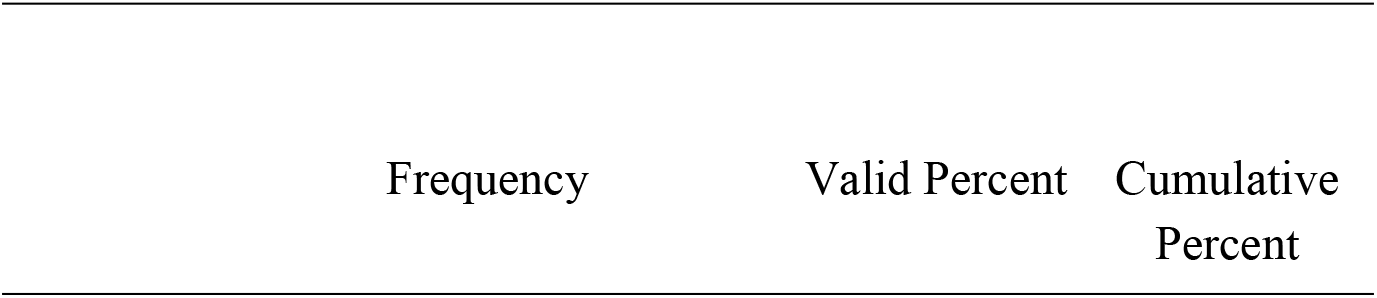
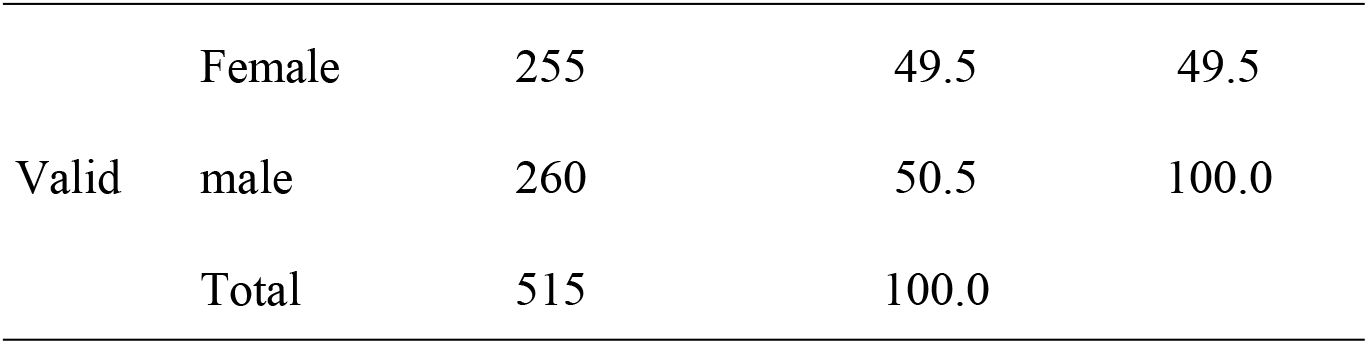
Descriptive statistics of Gender.

| Frequency | Valid Percent | Cumulative Percent |
| --- | --- | --- |

|  |  |  |  |  |
| --- | --- | --- | --- | --- |
|  | Female | 255 | 49.5 | 49.5 |
| Valid | male | 260 | 50.5 | 100.0 |
|  | Total | 515 | 100.0 |  |

**Table 1:**
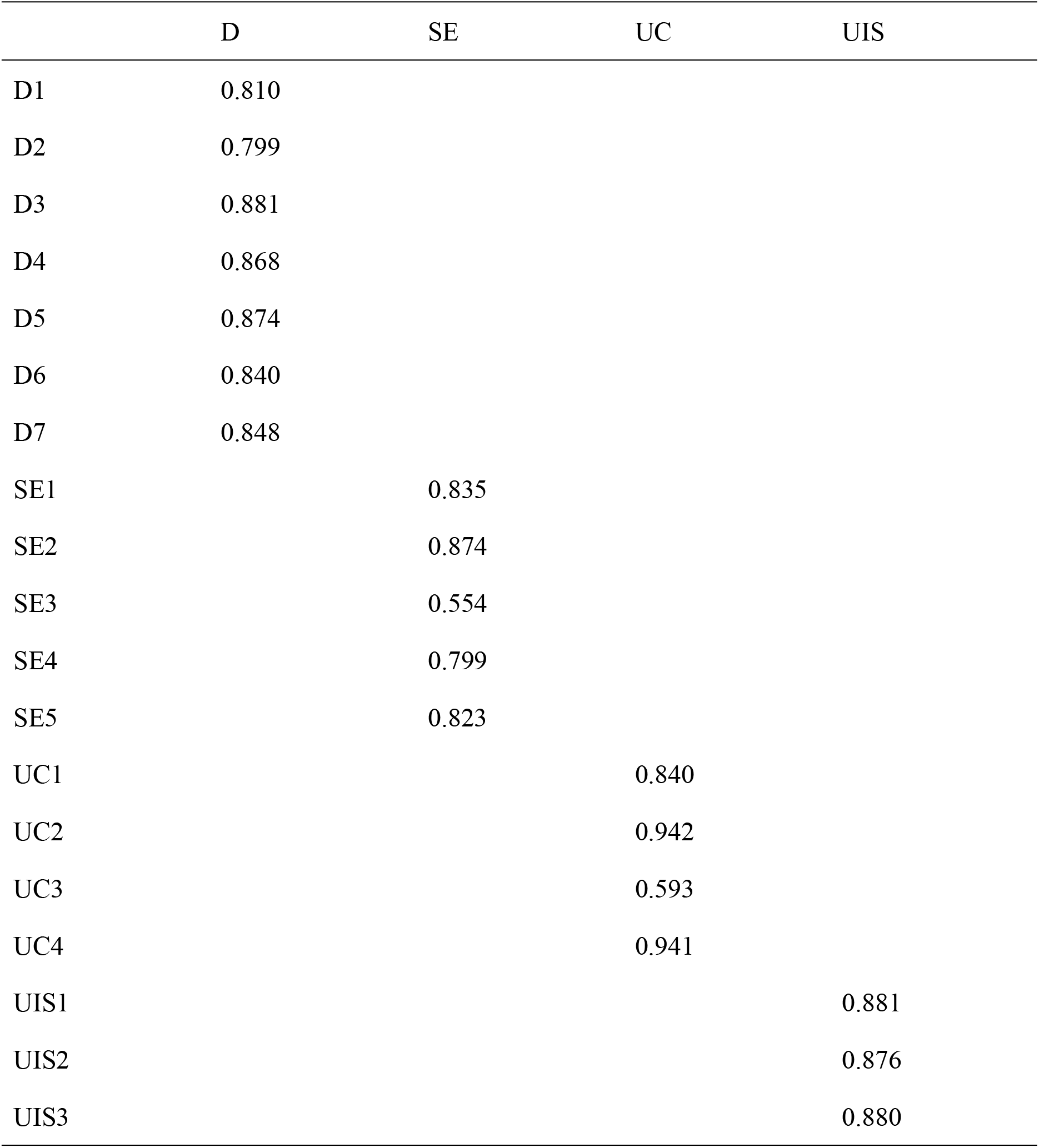
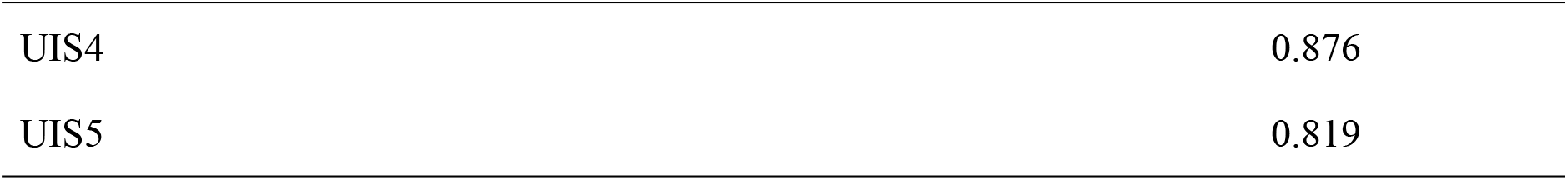
Factor Loadings.

We verified the complex model to check the mediation and see whether the moderator is changing the indirect effect (Figure 1). The usage of Instagram leads to depression through self-esteem and moderator upward comparison, which negatively influences the relationship between usage of Instagram and self-esteem (Hayes, 2018). Conditional mediation is when a model has one mediator and one moderator. An index term analysis is conducted when the moderator is continuous, and CoMe aims to assess the rate of change or relationship.

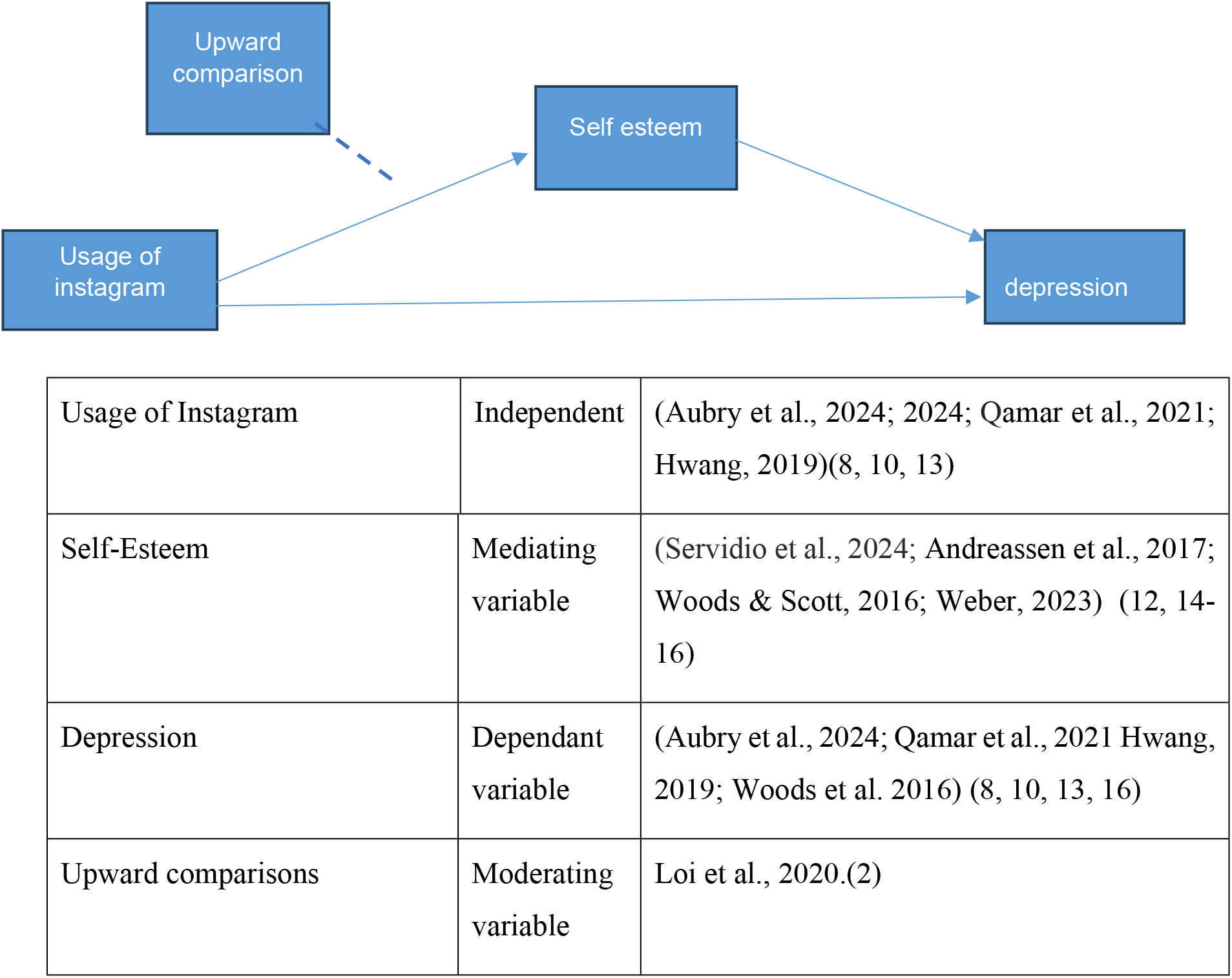

Figure 2 shows the conceptual and statistical model of Instagram and mental health.

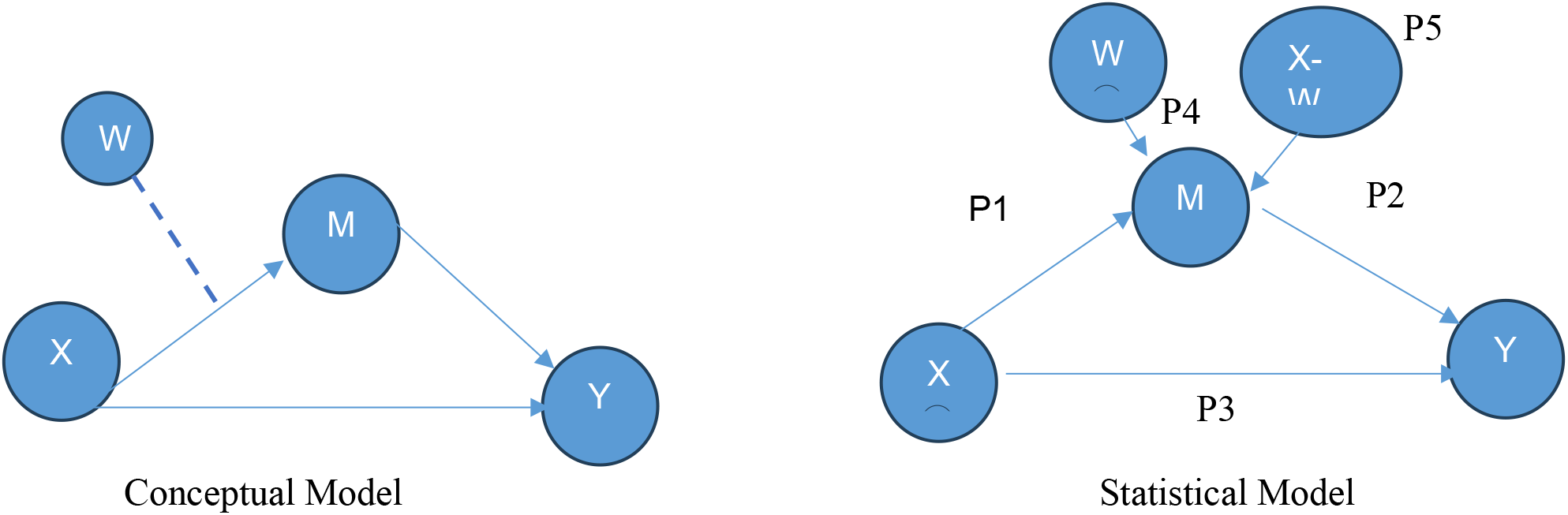

### Measurement Model

The constructs utilized in this study are evaluated through a measurement model executed in Smart PLS4. The assessment of quality criteria initiates with the factor loadings assigned to each item within the construct, subsequently followed by an evaluation of both construct reliability and construct validity. Table 1 presents the factor loading of the constructs. All factors exceed 0.50. All items in this study maintained a factor threshold of .50 (22). Therefore, no items have been removed. The values less than .50 could be removed but in this case, all items have greater than .50, see table 1.

### Reliability analysis

As noted by Meadows & Billington (2005), reliability refers to the degree of consistency and stability of the instrument. The primary approaches for assessing reliability are Cronbach’s alpha and composite reliability (CR). Table 2 shows the values of Cronbach’s reliability and Composite Reliability. The Cronbach’s alpha varied from 0.839 to 0.934, while composite reliability ranged from 0.869 to 0.934, indicating the reliability of all constructs under study and exceeding the threshold of 0.70 (23). Thus, the construct reliability has been established.

**Table 2.** Construct Reliability Analysis (Cronbach’s Alpha and Composite Reliability)

|  | Cronbach's alpha | Composite reliability |
| --- | --- | --- |
| D | 0.934 | 0.934 |
| SE | 0.839 | 0.869 |
| UC | 0.855 | 0.908 |
| UIS | 0.917 | 0.918 |

### Convergent Validity and Discriminant Validity

Convergent validity refers to the extent to which various methods of measuring the same concept yield consistent results. When the average variancer extracted value (greater than or equal to 0.50) convergent validity was established (24). The current study demonstrated that all constructs have AVE values exceeding 0.50, thereby confirming the establishment of convergent validity. Table 3 presents the AVE value for each construct.

**Table 3.**
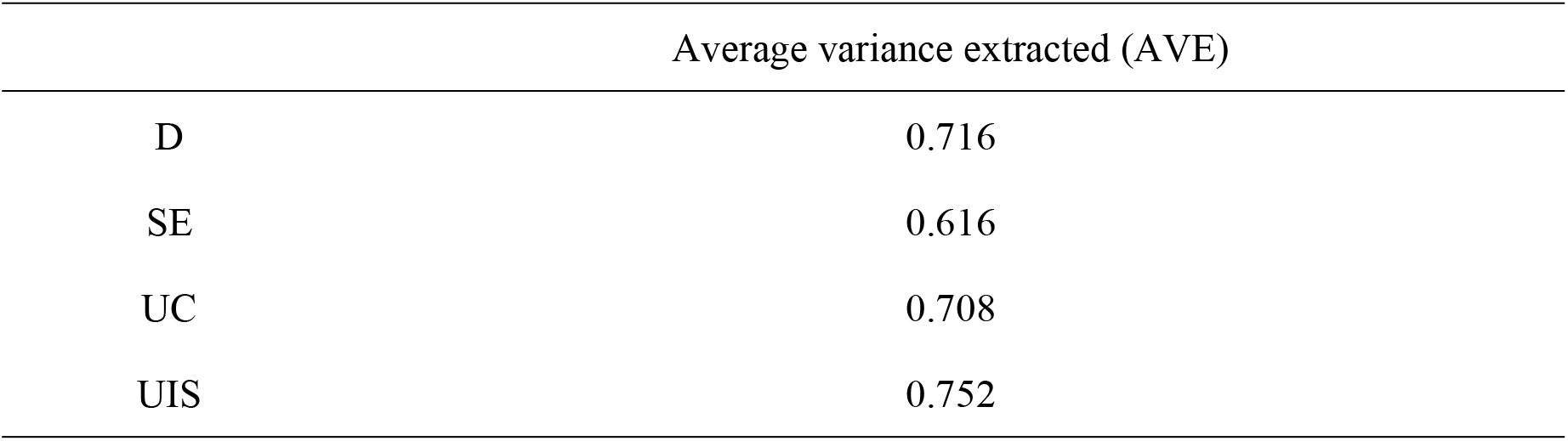
Construct Convergent Validity(AVE)

### Discriminant Validity

The extent to which measurement concepts are differentiated. The concept suggests that when two or more ideas are distinct, the valid measures for each should not exhibit a high degree of correlation (25). The researcher used the Fornell-Larker criteria for validating discriminant validity.

### Fornell Larker Criterion

Fornell and Larcker (24) state that criterion discriminant validity is confirmed when the square root of the Average Variance Extracted (AVE) for a construct exceeds its correlation with all other constructs. In this study, the square root of the Average Variance Extracted (AVE) for a construct exceeds its correlation with other constructs—Table 4. Therefore, discriminant validity is affirmed.

**Table 4:** Average Variance Extracted.

|  | D | SE | UC | UIS |
| --- | --- | --- | --- | --- |
| D | ***0.846*** |  |  |  |
| SE | -0.498 | ***0.785*** |  |  |
| UC | 0.726 | -0.639 | ***0.841*** |  |
| UIS | 0.329 | -0.625 | 0.358 | ***0.867*** |

***Note: bold and italics represent the square root of AVE***

### Structural Model

#### Conditional mediation analysis

We analyzed the relationship between Instagram usage and depression, focusing on the role of self-esteem as a mediator and the moderating effect of upward comparison. Researchers examine the influence of the moderator (UC) on the indirect pathway within a more intricate model. In the Conditional mediation analysis (CoMe)model, the independent variable (UIS - Usage of Instagram) influences the outcome variable Y (Depression D) through one mediator M (SE - self-esteem), and the relationship is conditioned by one moderator W, upward comparison UC (26). The p Value in table 5 indicated that all values are less .05 at 95% confidence interval.

**Table 5:** Moderated Mediation.

|  | Original sample (O) | Sample mean (M) | Standard deviation (STDEV) | T statistics (O/STDEV) | P values |
| --- | --- | --- | --- | --- | --- |
| UC -> SE -> D | 0.229 | 0.233 | 0.058 | 3.953 | 0.00 |
| UIS -> SE -> D | 0.29 | 0.289 | 0.069 | 4.178 | 0.00 |
| UC x UIS -> SE -> D | -0.035 | -0.034 | 0.016 | 2.138 | 0.016 |

Hence, *H1* of this study is accepted.

In the context of conditional mediation analysis, index term analysis is employed. The indices of P2 and P5 can be directly obtained from SmartPLS, as illustrated in Figure 2. The findings indicate that P2*P5 (or UC*UIS -> SE -> D) is statistically significant (p < .05). Given that the index “P2*P5 of UC is significant (β = -0.035), it can be concluded that the mediated effect of UIS on D through ES is negatively influenced by the moderator UC.

Index =-0.035 (.079 (UC*UIS->SE)* -0.439(SE->D))

### Reporting Moderated Mediation

Table 6 explains the direct relationship between constructs and answers RQ1. A notable negative correlation was identified between Instagram usage and self-esteem (β = -0.661, t = 5.091, p < .001), suggesting that increased use of Instagram is linked to diminished self-esteem. Self-esteem was found to be a significant predictor of depression (β = -0.439, t = 6.302, p < .001), indicating that lower self-esteem correlates with increased levels of depression. Instagram usage demonstrated a notable direct effect on depression (β = 0.343, t = 5.349, p < .001), indicating a positive association with depression that persists beyond the indirect impact.

**Table 6:** Direct Relationship.

|  | Original sample<br>(O) | T<br>statistics | P<br>values |
| --- | --- | --- | --- |
| Self-esteem -> Depression | -0.439 | 6.302 | 0.000 |
| Usage of Instagram -> Depression | 0.343 | 5.349 | 0.000 |
| Usage of Instagram -> Self-esteem | -0.661 | 5.091 | 0.000 |
| Upward comparison x Usage of Instagram -> Self-esteem | 0.079 | 2.281 | 0.011 |

***\*A T-statistic value greater than 1.645 means it is significant at .005***

The interaction between upward comparison and Instagram usage on self-esteem was found to be significant (β = 0.079, t = 2.281, p = .011), which suggests that the impact of Instagram usage on self-esteem is influenced by the degree of upward comparison. This indicates the existence of a moderation effect.

Table 7 answered RQ2. The indirect effect of Instagram on depression through self-esteem is significant at the .000 level. The analysis examined whether self-esteem serves as a mediator in the relationship between Instagram use and depression, assessing the significance of this indirect pathway. The findings indicated a significant indirect effect of Instagram use on depression through self-esteem (indirect effect = 0.290, t = 4.178, p < .001). This suggests that Instagram use contributes to increased depression, in part by diminishing self-esteem, which subsequently leads to higher levels of depression. Conversely, the direct effect of Instagram use on depression, after considering the influence of self-esteem, was not significant (direct effect = 0.052, t = 1.069, p = 0.143). The analysis confirmed that the indirect effect (Instagram → self-esteem → depression) is significant (p < .001), while the direct effect (Instagram → depression) is not significant (p = 0.143) when self-esteem is included in the model. Thus, the relationship between Instagram use and depression is fully mediated by self-esteem.

**Table 7:** Moderated Indirect Relationship.

|  | direct effect | indirect effect |
| --- | --- | --- |
| Usage of Instagram -> self-esteem -> depression | .052<br>(1.069) | .290<br>(4.178) |
| p value | 0.143 | 0.000 |

**Table 8:** Conditional indirect effect at different levels of the moderator (UC)

|  | Original<br>sample<br>(O) | Sample<br>mean<br>(M) | Standard<br>deviation<br>(STDEV) | T statistics<br>(O/STDEV) | P values |
| --- | --- | --- | --- | --- | --- |
| UIS → SE → D<br>conditional on UC at +1<br>SD | 0.116 | 0.118 | 0.034 | 3.452 | 0.000 |
| UIS → SE → D<br>conditional on UC at -1<br>SD | 0.201 | 0.203 | 0.036 | 5.527 | 0.000 |
| UIS → SE → D<br>conditional on UC at<br>Mean | 0.159 | 0.16 | 0.027 | 5.905 | 0.000 |

The table below indicates that the indirect path of usage of Instagram (UIS) leads to depression (D) through the mediator (SE), demonstrating a low value at a high level of the moderator (UC) = .0116. Conversely, the value of the indirect effect increases at a lower level of the moderator (UC) = .201. Although it is significant at all levels of moderation. The table indicates that Instagram (UIS) indirectly influences depression (D) through self-esteem (SE), which is impacted by upward comparison (UC). Instagram has a slight indirect influence on depression (.0116) when people compare themselves to others (high UC). The effect is larger (.201) when individuals make fewer comparisons (low UC). If people do not compare themselves to others, Instagram use might diminish self-esteem and cause depression. Although this indirect impact is present at all comparison levels, its intensity depends on the extent of the comparison.

## Discussion

This study examined a moderated mediation model to analyze the relationship between Instagram use (UIS) and depression (D), with self-esteem (SE) serving as a mediator and upward social comparison (UC) acting as a moderator. The findings indicate a nuanced relationship among these variables, offering valuable insights into the ways social media platforms such as Instagram can influence users’ mental wellbeing, especially concerning self-perceptions and tendencies toward comparison.

Our research demonstrates that self-esteem plays a significant mediating role in the relationship between Instagram usage and depression. Increased usage of Instagram correlated with decreased self-esteem (β= -0.661, p < .001), which subsequently predicted elevated levels of depression (β= -0.439, p < .001). This aligns with research indicating that regular interaction with social media platforms, particularly those focused on images such as Instagram, may have detrimental effects on self-esteem (Huang, 2009; Rose et al., 2014). The indirect relationship between Instagram usage and depression via self-esteem was found to be statistically significant (indirect effect = 0.290, t = 4.178, p < .001). In contrast, the direct effect was rendered non-significant upon the inclusion of the mediator (direct effect = 0.052, t = 1.069, p = 0.143). This pattern indicates complete mediation, which suggests that Instagram’s effect on depression is primarily mediated by its influence on users’ self-esteem.

The analysis revealed that upward comparison significantly interacted with Instagram usage in predicting self-esteem (β= 0.079, p = .011), thereby confirming the existence of a moderation effect. The conditional indirect effect of Instagram use on depression through self-esteem was found to be weaker at high levels of upward comparison (effect = 0.116 at +1 SD) and stronger at low levels of upward comparison (effect = 0.201 at -1 SD), while remaining significant across all levels. The index of moderated mediation was significant (β = -0.035, p = .016), indicating that the strength of the indirect path fluctuates according to the levels of upward comparison.

This finding may seem paradoxical initially; one might anticipate that individuals who often engage in upward comparison would experience a greater impact from Instagram use regarding self-esteem and depression. Our findings indicate a contrary perspective: the influence of Instagram usage on self-esteem, and consequently on depression, is more significant when upward comparison is minimal. A potential reason is that individuals strongly inclined to compare themselves to others may experience persistently low self-esteem, indicating that Instagram use does not significantly exacerbate their self-assessments. In their case, depression might be influenced by additional psychological factors not addressed in this study, including envy (27), anxiety about missing out (28), or feelings of social isolation (29).

On the other hand, individuals who engage less in upward comparison may not frequently evaluate their self-worth in everyday situations. However, when individuals engage extensively with Instagram, a platform characterized by idealized images, they may encounter a significant decline in self-esteem due to their lack of familiarity with such social comparisons. This abrupt change may clarify why the indirect impact of Instagram usage on depression is more pronounced among these individuals. The findings are consistent with the dual-pathway model of social media effects put forth (30), indicating that social comparison and self-esteem collaboratively influence the emotional outcomes associated with social media usage.

The notable indirect effect observed across all levels of upward comparison suggests that self-esteem consistently influences the relationship between Instagram use and depression. Nonetheless, the influence of this role fluctuates, underscoring the necessity of accounting for individual variations in social comparison tendencies when examining the effects of social media. Our findings align with previous research indicating that social media can adversely affect well-being by reducing self-esteem (20, 31).

Some studies have reported varying outcomes. Verduyn, Ybarra (32) posited that passive social media use, characterized by scrolling without interaction, results in an adverse effect due to upward comparison, in contrast to active use, which does not have the same effect. Our study did not distinguish between passive and active use, which could be considered a limitation. Furthermore, Tandoc Jr, Ferrucci (33) indicated that the use of Facebook is a predictor of depression, mainly through the mechanisms of social comparison and rumination. This reinforces the notion that the psychological effects of social media platforms are contingent upon context and shaped by individual traits and behaviors.

An alternative interpretation of our findings may be found in the concept of social comparison orientation (34) which denotes a predisposition to evaluate oneself in relation to others. Individuals with a strong inclination towards social comparison may experience a consistent impact from upward comparisons, while for others, Instagram could catalyze infrequent comparisons, resulting in heightened emotional reactions. It has been found that individuals evaluate their own worth by comparing themselves to others (35, 36). In the context of Instagram, such comparisons often involve idealized portrayals of lifestyles, appearances, and accomplishments, commonly leading to decreased self-esteem. This research advances the theoretical framework by illustrating that the impact of these social comparisons on self-esteem and depression is moderated by an individual’s inherent tendency to engage in upward comparisons.

### Implications

The findings have significant implications for mental wellbeing interventions and digital literacy programmes. Educating users, especially young adults, about the potential harmful effects of upward comparison and promoting mindful engagement with social media content can help lessen its negative impact on mental health. Psychological strategies such as mindfulness and self-compassion can be effectively taught to reduce social media-related comparison and boost self-esteem. Additionally, platform designers might explore features that minimize comparison triggers, such as concealing like counts or encouraging a variety of authentic content. It is essential for clinicians engaging with young clients to evaluate their social media usage and their inclination towards social comparison as a component of comprehensive mental health care.

### Limitations and Future Research

This study has certain limitations. The cross-sectional nature of the data limits the ability to establish causality. Future research should employ longitudinal or experimental designs to confirm the direction of effects. Additionally, distinguishing between different types of Instagram use (such as active versus passive) and expanding the model to include additional mediators (like envy, FoMO, and loneliness) may offer a more comprehensive understanding of the links between social media engagement and mental health outcomes.

## Conclusion

This study provides compelling evidence demonstrating that self-esteem functions as a full mediator in the relationship between Instagram usage and depression. Furthermore, this mediation process appears to be significantly influenced by upward social comparison, highlighting the complex interplay between online behaviors and psychological outcomes. These findings underscore the importance of considering individual differences in social comparison tendencies when assessing the psychological effects associated with social media platforms. As Instagram continues to play a prominent role in shaping self-perceptions and emotional well-being, it is essential for researchers and mental health practitioners to thoroughly understand these intricate pathways. Such insights can inform targeted interventions and guide the development of strategies aimed at mitigating adverse mental health effects related to social media engagement, ultimately promoting healthier online interactions and psychological resilience.

## Data Availability

Data can be provided as per request.

## Ethical approval

The study adhered to the principles outlined in the Helsinki Declaration and was approved by the relevant institutional review board as per letter number: FJWU/EC/2025/105 on May 19, 2025.

## Availability of data and material

The datasets generated during and/or analyzed during the current study are available from the corresponding author on reasonable request.

## Author Contribution

Conceptualization and Methodology: AQ and IA. Investigation and Data Curation: AQ. Formal Analysis: AQ, IA. Software and Visualization: AQ. Writing – Original Draft: AQ. Writing – Review & Editing: IA. Validation: AQ, IA.

## Acknowledgments

We are grateful to our interlocutors for their time and data.

## Consent to Participate and Publish

Informed consent was obtained from all participants to participate and to have their data published.

## Clinical trial

Not applicable.

